# Low booster uptake in cancer patients despite health benefits

**DOI:** 10.1101/2023.10.25.23297483

**Authors:** Jane C. Figueiredo, Julia Levy, So Yung Choi, Alexander M. Xu, Noah M. Merin, Omid Hamid, Tucker Lemos, Nathalie Nguyen, Maimoona Nadri, Alma Gonzalez, Simeon Mahov, Justin M. Darrah, Jun Gong, Ronald L. Paquette, Alain C. Mita, Robert A. Vescio, Sarah J. Salvy, Inderjit Mehmi, Andrew E. Hendifar, Ronald Natale, Warren G. Tourtellotte, V. Krishnan Ramanujan, Carissa A. Huynh, Kimia Sobhani, Karen L. Reckamp, Akil A. Merchant

## Abstract

Patients with cancer are at increased risk of death from COVID-19 and have reduced immune responses to SARS-CoV2 vaccines, necessitating regular boosters. We performed comprehensive chart reviews, surveys of patients attitudes, serology for SARS-CoV-2 antibodies and T-cell receptor (TCR) β sequencing for cellular responses on a cohort of 982 cancer patients receiving active cancer therapy accrued between November-3-2020 and Mar-31-2023. We found that 92·3% of patients received the primer vaccine, 70·8% received one monovalent booster, but only 30·1% received a bivalent booster. Booster uptake was lower under age 50, and among African American or Hispanic patients. Nearly all patients seroconverted after 2+ booster vaccinations (>99%) and improved cellular responses, demonstrating that repeated boosters could overcome poor response to vaccination. Receipt of booster vaccinations was associated with a lower risk of all-cause mortality (HR=0·61, P=0·024). Booster uptake in high-risk cancer patients remains low and strategies to encourage booster uptake are needed.

**Highlights:**

- COVID-19 booster vaccinations increase antibody levels and maintain T-cell responses against SARS-CoV-2 in patients receiving various anti-cancer therapies
- Booster vaccinations reduced all-cause mortality in patients
- A significant proportion of patients remain unboosted and strategies are needed to encourage patients to be up-to-date with vaccinations

## Introduction

Effective vaccines were developed, tested, and issued emergency use authorization (EUA) by the U.S. Food and Drug Administration (FDA), European Medical Agency and other regulatory agencies around the world in record time. Immunocompromised individuals were prioritized in vaccine rollout strategies;[1] however, none of the Phase III clinical trials included patients with cancer, fueling concerns about potential side effects and the robustness of their post-vaccine immune responses.[2] Data quickly emerged to reassure patients that available vaccines were safe and effective,[3] and subsequent studies reported that over 90% of patients with cancer have received at least one COVID-19 vaccine.[4] However, it is unclear how many are following recommendations for additional doses or “boosters.”

Booster vaccinations have been reported to counteract waning immune responses to vaccination and natural infection in healthy populations.[5] Several studies of the Omicron (B.1.1.529) subvariants reported heightened capacity for immune evasion,[6] and reduced vaccine effectiveness for first-generation monovalent COVID-19 vaccines.[7] Other reports suggest that additional doses of the monovalent mRNA vaccines,[8, 9] heterologous boosting,[10, 11] and the bivalent mRNA vaccines provide additional immune protection to SARS-CoV-2 B.1.1.529 infection and severe COVID-19 illness.[12]

For patients with cancer, evidence suggests that primer vaccinations elicit lower antibody titers compared to healthy individuals, albeit dependent on disease state, co-morbidities, and anticancer treatment status.[13–18] Subsequent booster vaccinations have been reported to restore and increase antibody titers in patients similar to healthy controls,[11, 18, 19] except for patients receiving B-cell-depleting therapies and chemotherapies.[11, 19] However, even seronegative individuals appear capable of mounting T-cell responses,[14, 20] and some data suggest that T-cell responses are more robust over time compared to antibody levels.[21]

Changes in vaccine availability and the evolving nature of the science and public health recommendations for additional doses have likely impacted the psychosocial wellbeing of patients. During the pandemic, there was an increase in depression and anxiety rates among patients with cancer, which was exacerbated by social distancing.[22] Disruptions in cancer care services were also reported to contribute to higher levels of anxiety, loneliness and social isolation among patients.[22, 23]

There are unanswered questions about changes in the mental, social, and immunologic health of patients with cancer as we enter into the endemic phase of SARS-CoV-2. In this follow-up study from the SeroNet-CORALE Cancer Cohort Study, we report on vaccine uptake patterns, and examine humoral and cellular immune responses, psychosocial health, and all-cause mortality.

## Results

### Patient characteristics and attitudes on SARS-CoV-2 vaccines

We enrolled 986 patients with either solid (48·7%) or hematologic (51·3%) malignancies (median age=63·0 [IQR, 52·0-70·0] years; 48·6% women, **Table 1**, **Figure S1**). Less than 8% of patients refused vaccination, and <2·0% of survey responses reported being against vaccination at any timepoint during follow-up (**Figure S2**). Patients who agreed that vaccines may not be safe were less likely to be up-to-date with vaccinations at any timepoint (range: 11·8–40·0% v. 4·3–6·7%). Fear of adverse events has remained a common concern, but has decreased over time (46·2% from Jan to Aug–2021 v. 42·7% from Oct–2022 to Mar–2023).

**Table 1:** Characteristics of the study cohort.

|  | Primer |  | 1<sup>st</sup> booster (monovalent) |  | ≥2<sup>nd</sup> booster (monovalent) |  | Bivalent booster |  |
| --- | --- | --- | --- | --- | --- | --- | --- | --- |
|  | Solid<br>(n=444) | Hematologic<br>(n=466) | Solid<br>(n=347) | Hematologic<br>(n=351) | Solid<br>(n=156) | Hematologic<br>(n=181) | Solid<br>(n=152) | Hematologic<br>(n=145) |
| **Vaccinated proportion (%)** | 92.5% | 92.1% | 78.2% | 75.3% | 45.0% | 51.6% | 31.7% | 28.7% |
| **Age at enrollment, median (IQR) years** | 64.0 (55.0, 72.0) | 62.0 (51.0, 69.0) | 65 (56.0, 73.0) | 63.0 (53.0, 70.0) | 66.0 (59.8, 74.0) | 66.0 (56.0, 71.0) | 66.5 (60.0, 74.0) | 63.0 (54.0, 71.0) |
| **Age at diagnosis, n** | 416 | 445 | 322 | 333 | 145 | 174 | 137 | 139 |
| **Age at diagnosis, median (IQR) years** | 62.0 (52.0, 70.0) | 57.0 (47.0, 66.0) | 62.0 (52.0, 70.0) | 58.0 (50.0, 66.0) | 64.0 (55.0, 72.0) | 60.0 (52.0, 68.8) | 64.0 (55.0, 72.0) | 59.0 (51.0, 69.0) |
| **Female sex (n, %)** | 243 (54.7%) | 197 (42.3%) | 196 (56.5%) | 148 (42.2%) | 92 (59.0%) | 70 (38.7%) | 93 (61.2%) | 65 (44.8%) |
| **Race and ethnicity (n, %)** |  |  |  |  |  |  |  |  |
| Non-Hispanic White | 273 (62.0%) | 235 (50.5%) | 213 (61.9%) | 193 (55.1%) | 104 (67.1%) | 111 (61.3%) | 93 (61.6%) | 90 (62.1%) |
| Hispanic | 61 (13.9%) | 126 (27.1%) | 47 (13.7%) | 75 (21.4%) | 15 (9.7%) | 24 (13.3%) | 17 (11.3%) | 22 (15.2%) |
| Non-Hispanic Black | 29 (6.6%) | 54 (11.6%) | 22 (6.4%) | 42 (12.0%) | 7 (4.5%) | 26 (14.4%) | 14 (9.3%) | 16 (11.0%) |
| Asian | 44 (10.0%) | 34 (7.3%) | 37 (10.8%) | 28 (8.0%) | 19 (12.3%) | 16 (8.8%) | 14 (9.3%) | 15 (10.3%) |
| Other | 33 (7.5%) | 16 (3.4%) | 25 (7.3%) | 12 (3.4%) | 10 (6.5%) | 4 (2.2%) | 13 (8.6%) | 2 (1.4%) |
| **Anti-cancer treatment\* (n, %)** |  |  |  |  |  |  |  |  |
| Chemotherapy | 110 (24.8%) | 119 (25.5%) | 104 (30.0%) | 106 (30.2%) | 57 (36.5%) | 44 (24.3%) | 59 (38.8%) | 48 (33.1%) |
| B-cell therapy | 2 (0.5%) | 148 (31.8%) | 2 (0.6%) | 132 (37.6%) | 1 (0.6%) | 67 (37.0%) | 1 (0.7%) | 64 (44.1%) |
| Immune checkpoint inhibitors | 118 (26.6%) | 7 (1.5%) | 88 (25.4%) | 5 (1.4%) | 37 (23.7%) | 3 (1.7%) | 35 (23.0%) | 0 (0.0%) |
| Targeted therapy | 53 (11.9%) | 115 (24.7%) | 59 (17.0%) | 103 (29.3%) | 34 (21.8%) | 58 (32.0%) | 30 (19.7%) | 60 (41.4%) |
| Hormonal therapy | 35 (7.9%) | 10 (2.1%) | 31 (8.9%) | 5 (1.4%) | 19 (12.2%) | 1 (0.6%) | 17 (11.2%) | 2 (1.4%) |
| Stem cell transplant (allogeneic) | 0 (0.0%) | 43 (9.2%) | 0 (0.0%) | 36 (10.3%) | 0 (0.0%) | 22 (12.2%) | 0 (0.0%) | 17 (11.7%) |
| Stem cell transplant (autologous) | 0 (0.0%) | 28 (6.0%) | 0 (0.0%) | 25 (7.1%) | 0 (0.0%) | 17 (9.4%) | 0 (0.0%) | 8 (5.5%) |
| **ECOG Status (n, %)** |  |  |  |  |  |  |  |  |
| 0-1 | 241 (93.8%) | 289 (94.4%) | 243 (95.3%) | 267 (94.3%) | 130 (94.9%) | 163 (97.0%) | 129 (94.2%) | 129 (91.5%) |
| 2-4 | 16 (6.2%) | 17 (5.6%) | 12 (4.7%) | 16 (5.7%) | 7 (5.1%) | 5 (3.0%) | 8 (5.8%) | 12 (8.5%) |
| **Vaccine Type (n, %)** |  |  |  |  |  |  |  |  |
| Ad26.COV2.S | 19 (4.3%) | 18 (3.9%) | 4 (1.2%) | 3 (0.9%) | 0 (0.0%) | 1 (0.6%) | 84 (55.3%) | 84 (57.9%) |
| BNT162b2 | 224 (50.5%) | 268 (57.5%) | 166 (47.8%) | 206 (58.7%) | 68 (43.6%) | 99 (55.0%) | 68 (44.7%) | 61 (42.1%) |
| mRNA-1273 | 201 (45.3%) | 180 (38.6%) | 177 (51.0%) | 142 (40.5%) | 88 (56.4%) | 80 (44.4%) | 40 (26.3%) | 42 (29.0%) |
| **Heterologous boosting (n, %)** | -- | -- | 29 (8.4%) | 26 (7.4%) | 26 (16.7%) | 28 (15.6%) | 39 (25.7%) | 65 (44.8%) |
| **SARS-CoV-2 infection prior to vaccination (n, %)** | 19 (4.3%) | 32 (6.9%) | 32 (9.2%) | 51 (14.5%) | 18 (11.5%) | 38 (21.0%) | 0 (0.0%) | 1 (1.2%) |
| **Serology available (n, %)** | 161 (36.3%) | 243 (52.1%) | 174 (50.1%) | 215 (61.3%) | 83 (53.2%) | 120 (66.3%) | 69 (45.4%) | 84 (57.9%) |
| **No seroconversion after vaccination** | 5 (3.1%) | 38 (15.6%) | 1 (0.6%) | 12 (5.6%) | 1 (1.2%) | 1 (0.8%) | 0 (0.0%) | 1 (1.2%) |

\*Up to 3 months prior to the time point of vaccination indicated in the column

### SARS-CoV-2 vaccine booster uptake

Following FDA EUA recommendations for the 1^st^ monovalent booster in Aug–2021, a total of 698 patients (49·7% with solid tumors and 50·3% with hematologic malignancies) were boosted with a monovalent booster with a median time from first dose of 238·5 days (IQR=207·2–280) days. Most patients received a homologous first booster (94·6% BNT/BNT-primed with BNT 1^st^ monovalent booster; 93·6% m1273/m1273-primed with m1273 1^st^ monovalent booster), but heterologous boosting became more frequent for subsequent doses (only 84·0% of patients receiving 2^nd^ homologous booster which dropped to 62·5% for the 3^rd^ booster).

Less than 35% of patients received two monovalent booster doses (median of 196 days from the 1^st^ monovalent booster) and only 30·1% received at least one bivalent booster (95·6% after receiving at least one monovalent, **Figure 1 Panel A**). When we examined vaccine uptake by age groups and self-identified race and ethnicity, the lowest uptake was among patients under 50 years of age compared to those over age 70 (**Figure 1 Panel B**), and for individuals who self-identified as either Black or Hispanic compared to non-Hispanic White (**Figure 1 Panel C**).

**Figure 1:**
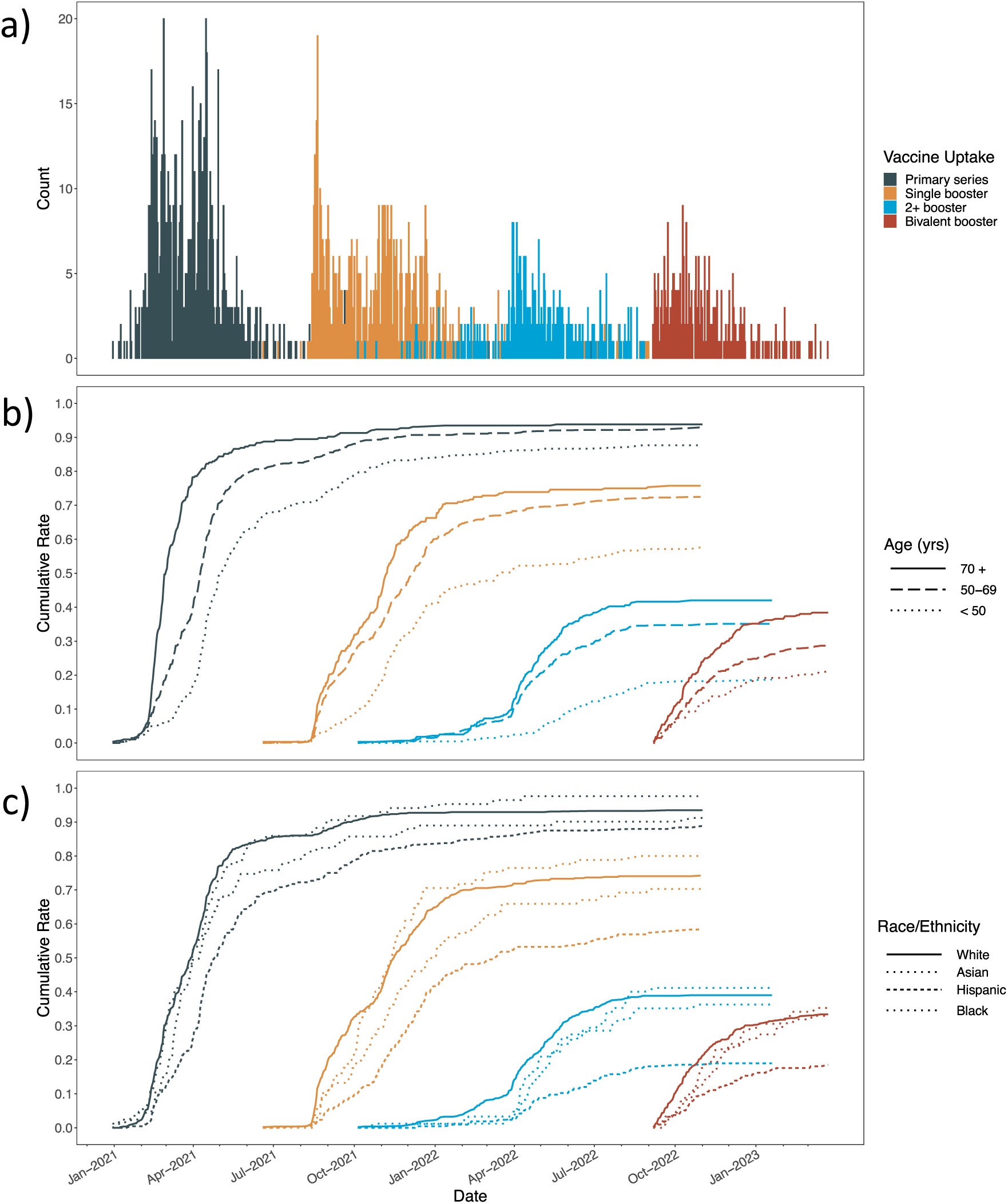
Dynamics of SARS-CoV-2 vaccine uptake after primer and booster vaccinations in patients with cancer. **Panel A.** Plot showing number of patients with cancer who received the primer series (dark blue), 1^st^ booster (orange), 2^nd^ or more boosters (light blue) and bivalent booster (red) over time. **Panel B.** Plot showing cumulative number of patients with cancer who received the primer series (dark blue), 1^st^ booster (orange), 2^nd^ or more boosters (light blue) and bivalent booster (red) over time by age categories (< 50, 50-59, 70+) (p-values for difference in vaccine uptake by age: <0.0001 (primer series), <0.0001 (1^st^ booster), <0.0001 (2^nd^+ booster), <0.0001 (bivalent booster). **Panel C.** Plot showing cumulative number of patients with cancer who received the primer series (dark blue), 1^st^ booster (orange), 2^nd^ or more boosters (light blue) and bivalent booster (red) over time by self-reported race and ethnicity (non-Hispanic White, non-Hispanic Black, Hispanic/Latino, Asian, Other) (p-value for difference in vaccine uptake by race and ethnicity: <0.0001 (primer series), <0.0001 (1^st^ booster), <0.0001 (2^nd^+ booster), 0.0003 (bivalent booster).

### Humoral immune responses post-vaccination

For the primer series, median peak IgG(S-RBD) antibody levels post-vaccination (available in 44·4% of patients) differed by demographic and clinical characteristics. Peak antibody levels were highest in patients <50 years of age (p<0·001), patients who self-identified as Hispanic (p=0·005), patients with a prior SARS-CoV-2 infection (p<0·001), and patients receiving B-cell therapies (p<0·001, **Figure 2 Panel A**). 10·6% of patients (88·4% hematologic malignancies) did not seroconvert after priming.

**Figure 2:**
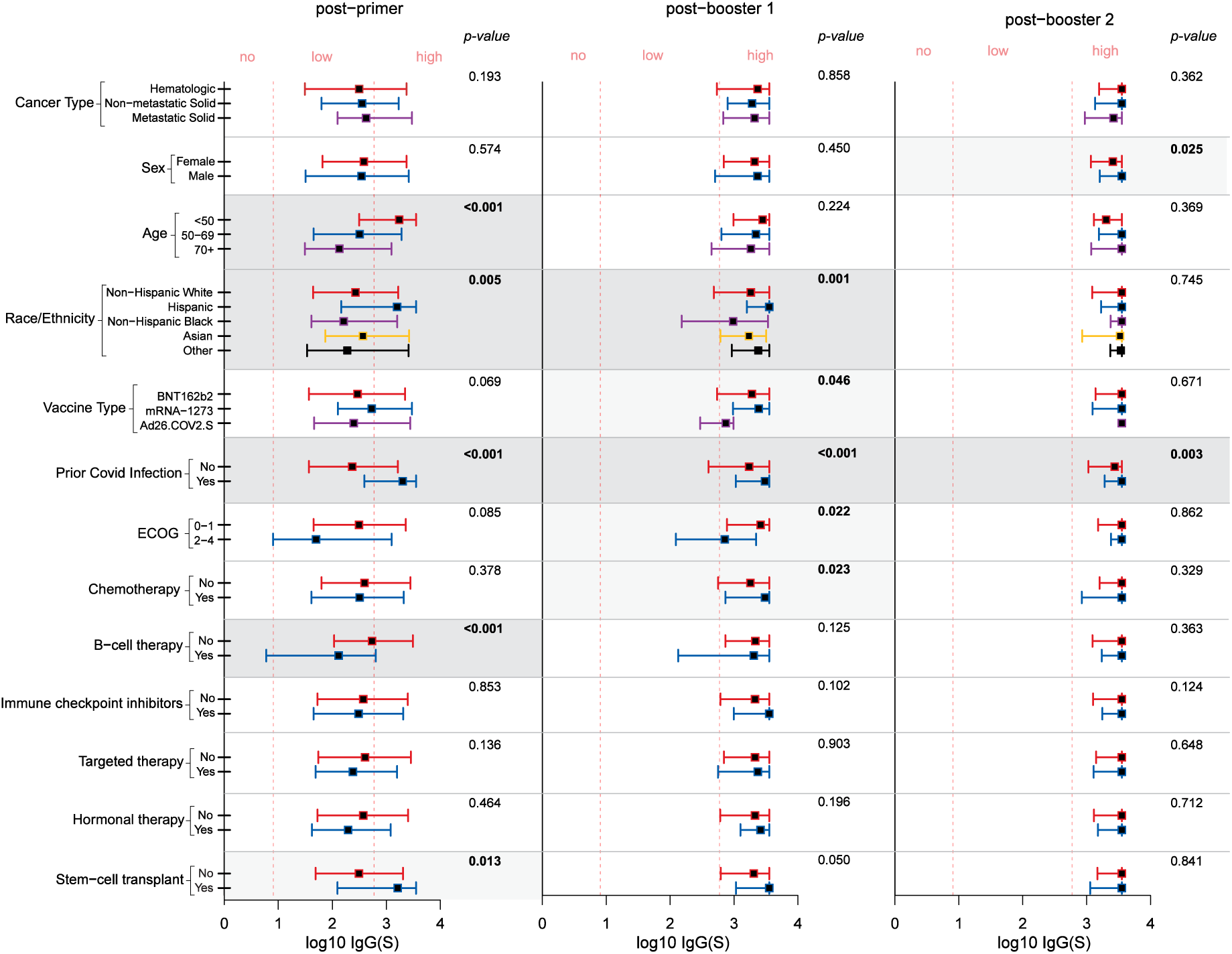
Box and whisker plots showing peak IgG (S-RBD) antibody levels after primer (n=404), post-monovalent booster #1 (n=389) and post-monovalent booster #2 (n=203) among patients with cancer by selected clinical and vaccine characteristics. P-values less than 0.05 are bolded.

Median peak IgG(S-RBD) levels increased with booster doses from 372 (IQR=51–2,475) BAU/ml to 2,150 (IQR=639–3,550) BAU/ml after one booster and to 3,550 (IQR=1,324–3,550) BAU/ml after two boosters. Factors related to antibody levels after one booster dose were: race/ethnicity (p=0·001), vaccine type (p=0·046), prior SARS-CoV-2 infection (p<0·001), ECOG status (p=0·022) and chemotherapy use (p=0·023, **Figure 2 Panel B**). Among those with 2+ boosters, nearly all reached the threshold of IgG(S-RBD) antibody levels; male patients and those with a prior COVID infection had higher peak levels (**Figure 2 Panel C**). Rates of non-seroconversion after boosting were: 3·3% after one monovalent dose and 1·0% after two monovalent doses.

### Cellular immune responses post-vaccination

Following primer vaccination, clonal breadth was lower among the patients who received B-cell therapy compared to those that did not (p=0·025); and among those receiving BNT and Ad26 compared to m1273 (p<0·001, **Figure S4, Panel A**). In contrast, clonal depth response was lower not only in patients receiving B-cell therapy (p<0·001) and those receiving BNT and Ad26 (p=0·015), but also in patients with hematological malignancies vs. solid tumors (p<0·001), male patients vs female (p<0·001), by age (p=0·031) and by race and ethnicity (p=0·034). (**Figure S4, Panel B**). Upon receiving one monovalent booster, these significant differences were not observed, other than male patients continuing to have lower clonal depth compared to female patients (p=0·015) (**Figure S4, Panels C and D**). Among patients who were seronegative after priming, 18·9% exhibited a T-cell response using the T-detect metric. These patients were all hematologic malignancies with most receiving B-cell targeted therapies. Among all patients after priming, we observed a modest correlation was between IgG(S) and breadth, and IgG(S) and depth [Spearman’s rho=0.602 and 0.595, p-values<0.001, respectively].

### Symptoms post-vaccination

Among vaccinated patients completing a post-vaccination survey, symptoms were highest for the second dose and lowest for the third dose. Symptoms included site pain (59·5%, 48·1%, 9·1%), fatigue (39·2%, 48·1%, 4·5%), headaches or dizziness (15·2%, 16·0%, 6·8%), fever (19·0%, 37·7%, 11·4%) and muscle aches (6·3%, 11·3%, 4·5%) after dose 1, 2 and 3, respectively (**Figure S3**). No serious toxicities attributable to vaccination were observed in the cohort.

### Patient-reported outcomes

Self-reported anxiety levels decreased from 50·7 (SD=9·4) during the period from Jan-2021 to Aug-2021 to 48·1 (SD=8·5) from Oct–2022 to Mar–2023 in all patient groups regardless of their infection and vaccination status (p=0·007, **Figure 3**). Social support as measured by its three constructs – informational, instrumental and emotional – also improved over time [Jan–2021 to Aug–2021: mean T-scores=53·9 (SD=11·1), 54·7 (SD=9·8), 54·9 (SD=8·5) and Sept–2021 to Mar–2022: mean T-scores=57·7 (SD=10·3), 57·7 (SD=8·1), 57·5 (SD=7·7), respectively]. Self-reported loneliness also decreased after the first booster was introduced (Jan–2021 to Aug–2021: 4·0 (SD=1·4) and Sept–2021 to Mar–2022: 3·8 (SD=1·5), p=0·042). However, depression levels remained constant over time.

**Figure 3:**
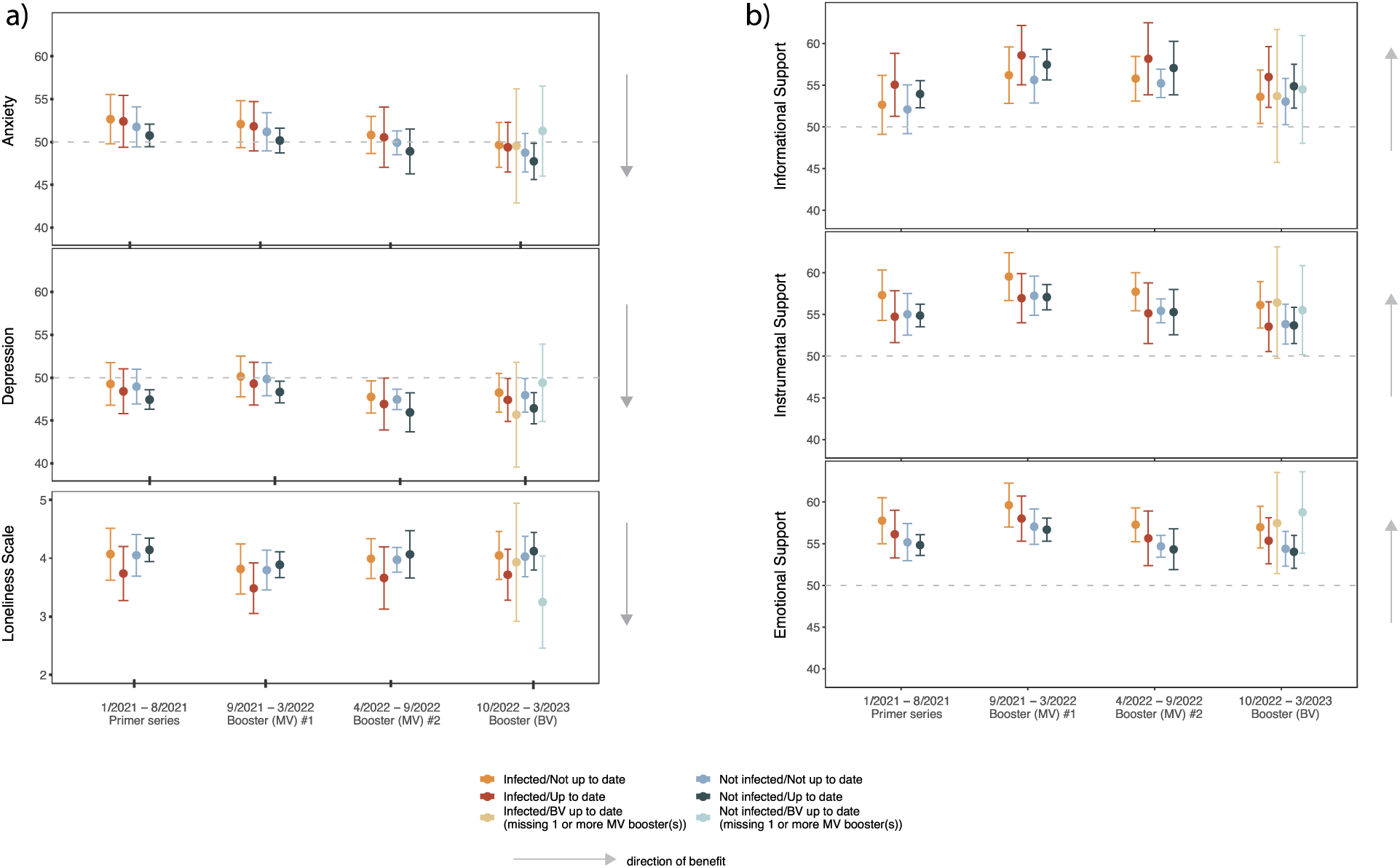
Change in six self-reported HR-QOL measures over the course of the pandemic defined by four time periods: (1) Jan-2021 through Aug-2021 when the primer series was recommended/available; (2) Sept-2021 through Mar-2022 when the first monovalent (MV) booster was recommended/available; (3) April-2022 through Sept-2022 when the second MV booster was recommended/available; and (4) Oct-2022 through Mar-2023 when a bivalent booster was recommended/available. **Panel A** show scores for anxiety, depression and loneliness; and **Panel B** show scores for informational support, instrumental support and emotional support. Scores for anxiety, depression and support were converted to a T-score, which is used to compare sample scores versus the general population (mean=50, SD=10). Higher T-scores correspond to higher levels. For the loneliness scale, scores were summed and ranged from 3 (least) to 9 (most lonely). The arrow depicts the direction of benefit. For each of the four time periods, least-squares means and 95% confidence limits were plotted for patients separately by their infection and vaccination status at that time.

### Predictors of SARS-CoV-2 infection and all-cause mortality

Overall, 40·4% of patients in this patient cohort were infected either prior to (3·5%) or after (33·2%) vaccination including multiple infections. Multiple infections were reported by 19.2% of patients, and 3·7% of patients had infections both prior to and after vaccination. The majority of breakthrough infections was after Dec–2021 when Omicron became the dominant lineage in the county. In multivariable Cox model, B–cell treatment (HR=1·46, 95% CI=1·17–1·82, p=0·001), ECOG status (HR=0·69, 95% CI=0·48–1·00, p=0·0497), and prior covid infection(s) (HR=3·51, 95% CI=2·85–4·34, p<0·001) were significant risk factors for infection (**Table S8**). Hospitalization and ICU admissions for COVID-19 after the emergence of Omicron were less common than before (27·9% v. 85·3% and 5·9% v. 19·2%, respectively).

In multivariable Cox model, all-cause mortality was associated with male sex (HR=1·72, 95% CI=1·14–2·57, p=0·009), receipt of chemotherapy (HR=1·76, 95% CI=1·18–2·63, p=0·006), ECOG status of 2 to 4 (HR=2·27, 95% CI=1·27–4·06, p=0·006) and not receiving a booster dose (HR=0·61, 95% CI=0·40–0·94, p=0·024, **Table 2**). By cancer type, receiving a booster dose was significantly associated with reduced mortality in adjusted models for those with hematologic malignancies (HR=0.48, 95% CI=0·24–0·94, p=0.032), but not among individuals with solid malignancies (HR=0.82, 95% CI=0·45–1·51, p=0.528).

**Table 2:**
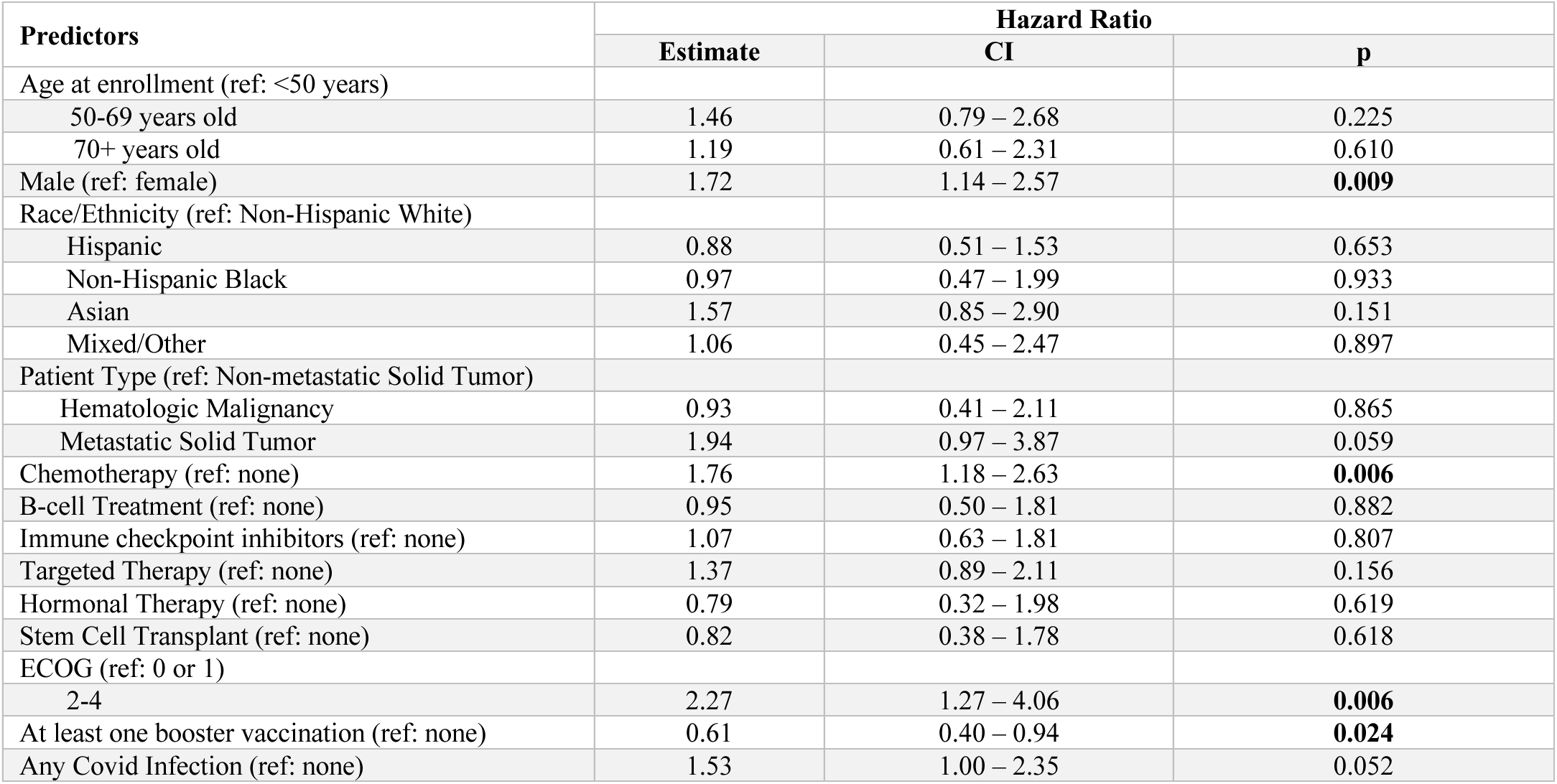
Multivariable model for all-cause mortality among patients with cancer in this cohort.

| Predictors | Hazard Ratio |  |  |
| --- | --- | --- | --- |
|  | Estimate | CI | p |
| Age at enrollment (ref: <50 years) |  |  |  |
| 50-69 years old | 1.46 | 0.79 – 2.68 | 0.225 |
| 70+ years old | 1.19 | 0.61 – 2.31 | 0.610 |
| Male (ref: female) | 1.72 | 1.14 – 2.57 | **0.009** |
| Race/Ethnicity (ref: Non-Hispanic White) |  |  |  |
| Hispanic | 0.88 | 0.51 – 1.53 | 0.653 |
| Non-Hispanic Black | 0.97 | 0.47 – 1.99 | 0.933 |
| Asian | 1.57 | 0.85 – 2.90 | 0.151 |
| Mixed/Other | 1.06 | 0.45 – 2.47 | 0.897 |
| Patient Type (ref: Non-metastatic Solid Tumor) |  |  |  |
| Hematologic Malignancy | 0.93 | 0.41 – 2.11 | 0.865 |
| Metastatic Solid Tumor | 1.94 | 0.97 – 3.87 | 0.059 |
| Chemotherapy (ref: none) | 1.76 | 1.18 – 2.63 | **0.006** |
| B-cell Treatment (ref: none) | 0.95 | 0.50 – 1.81 | 0.882 |
| Immune checkpoint inhibitors (ref: none) | 1.07 | 0.63 – 1.81 | 0.807 |
| Targeted Therapy (ref: none) | 1.37 | 0.89 – 2.11 | 0.156 |
| Hormonal Therapy (ref: none) | 0.79 | 0.32 – 1.98 | 0.619 |
| Stem Cell Transplant (ref: none) | 0.82 | 0.38 – 1.78 | 0.618 |
| ECOG (ref: 0 or 1) |  |  |  |
| 2-4 | 2.27 | 1.27 – 4.06 | **0.006** |
| At least one booster vaccination (ref: none) | 0.61 | 0.40 – 0.94 | **0.024** |
| Any Covid Infection (ref: none) | 1.53 | 1.00 – 2.35 | 0.052 |

## Discussion

In this cohort study, nearly all patients with cancer who received two or more boosters reached the threshold level indicative of seroconversion. Booster vaccinations did not appear to significantly reduce risk of infection and over 40% of patients in our cohort became infected with SARS-CoV-2 after vaccination, the majority after the emergence of Omicron. However, the likelihood of severe illness and COVID-related death were exceedingly low. There was also a significantly reduced risk of all-cause mortality associated with booster vaccinations after adjustment for known demographic and clinical variables. Despite this evidence of protection, booster uptake was low in this cohort of patients, with 21·9% who remain unboosted with either a monovalent or bivalent booster.

Booster uptake did not appear to be linked to vaccine hesitancy in this cohort. In our first report early in the pandemic, we observed higher hesitancy related to vaccine safety and adverse events especially among female compared to male patients.[24] Other studies examining vaccine hesitancy among patients with cancer from 2020 to 2021 also reported patients were concerned about vaccine safety, adverse effects, and efficacy due to the rapid development and novelty of mRNA vaccines,[25, 26] with only 49% of patients reporting that they had discussed vaccination with their oncologist.[26] In this updated report, we show that patients remain concerned about vaccine safety, efficacy and occurrence of adverse events, but this proportion has decreased with time and the availability of booster vaccinations.

The importance of additional booster doses in maintaining immune protection against SARS-CoV-2 has been documented in this cohort and other studies.[18, 19, 27] Although vaccination is associated with robust serological response in most cancer patients following vaccination, the durability of this response is reduced in cancer patients.[13] Booster vaccinations appears to increase titers beyond levels initially elicited with the primer series, even among patients with initially low or no antibody titers after two vaccine doses in the primer series. Serial boosters may continue to strengthen and help maintain the humoral immune response in the vast majority of patients with cancer under active treatment. Still some patients, including those receiving anti-CD20 therapy, CD19-directed chimeric antigen receptor T-cell therapy and ibrutinib,[11],[19, 28] may require alternative protective measures such as recombinant antibodies or high titer intravenous immunoglobulin.

In addition to the humoral cellular response, we measured the T cell response post-vaccination. While TCR response as measured by breadth/depth is influenced by HLA type,[29] the overall trends indicated that TCR response was correlated with humoral response, including for those with hematologic malignancies. Previous studies have shown T-cell vaccination responses in 46– 88% of patients with solid tumors and 45–75% of patients with hematological cancers.[14, 30] In patients with cancer, reduced T-cell responses to vaccination have been associated with Bruton tyrosine kinase inhibitors and steroid use.[30],[31] Immune checkpoint inhibitors have been found to be associated with increased T-cell immunity,[32] although it remains to be seen how other explicitly T-cell-involved therapies and malignancies such as bispecific T-cell engagers for hematologic malignancies or T cell lymphomas interact with vaccination. Additional factors to consider for T cell immunity are the effects of exhaustion or cancer-associated T-cell anergy,[33] which can prevent existing clones from expanding. This phenomenon is reflected by differential clonal breadth versus depth responses, for instance in hematological cancers, where patients have similar clonal breadth response as solid tumor patients but poor clonal depth, indicating a lack of clonal expansion. In our data, older patients, male patients, and B-cell therapy patients all exhibit this same disparity, which is abrogated in all groups after boosters, except in male patients. Thus, while boosters do not appear to significantly increase T-cell immunity, they may address differences seen after primer series. Additional functional studies of T cells cancer patients after boosters are needed to better coorelate TCR clonal studies with direct functional assays.

We also noted improvement in self-reported anxiety levels in patients in this cohort over the last two years of the pandemic regardless of their vaccine status or infection history. However, depression did not significantly change over time, and remained lower than the average cancer population. The World Health Organization reported that the COVID-19 pandemic triggered a 25% increase in the prevalence of anxiety and depression worldwide.[34] In patients with cancer, previous studies reported heightened anxiety and depression as a results of the uncertainty of the pandemic and associated delays in cancer diagnosis or treatment.[22, 23] Patients also expressed fear and worry about the implications of their immunodeficiences and the negative consequences of contracting COVID-19 infection.[23] Our data would suggest improvement in pandemic-induced anxiety, but further data is needed to confirm this observation.

We further evaluated changes in three types of social support among patients in this cohort: instrumental, emotional and informational. We observed significant improvement in all three types of social support after the first booster was introduced in Aug-2021, but no substantive changes since that time. We also observed lower levels of loneliness once boosters were introduced compared to early in the pandemic. Other reports highlighted loneliness as a potential concern among immunocompromised patients who were more likely to practice recommended preventive social distancing measures than adults without cancer,[35] leading to increased social isolation.[22] However, in recent years as the COVID-19 shifted from pandemic to endemic, more guidance on vaccination schedules became available in addition to treatment options for illness, and accordingly greater reassurance that has increased social interactions.

There are several limitations in this study. Observational studies are subject to potential selection biases, missing data and other unmeasured confounders. In this cohort study, we did not have complete serological or cellular immune data at all timepoints. Furthemore, patients who are seropositive would be expected to have a T cell response, but our finding of discordant results could suggest a limitation of the T detect assay. Notably, the percentage of discordance between serologic response and TCR response differed by race and ethnicity, suggesting that perhaps differences in HLA could play a part in this discordance. Our sample size limitations also precluded analysis of certain subgroups of interest.

In summary, this study provides evidence that booster vaccination improved immune responses and reduced risk of mortality in cancer patients. Additional doses of COVID-19 vaccines are recommended for immunocompromised individuals, but a substantial proportion of patients are not receiving boosters. Avoiding pandemic-fatigue and simplifying an immunization schedule may encourage more patients to keep up-to-date with vaccinations.

## Methods

### Study Design and Participants

The U.S. National Cancer Institute-funded Serological Sciences Network (SeroNet)-Coronavirus Risk Associations and Longitudinal Evaluation (CORALE)[13, 24] study is a longitudinal cohort including cancer patients over age 18 with histologically confirmed solid or hematological malignancies. Patients were recruited in the clinical setting from Nov-3-2020 to Mar-31-2023. To focus on cancer patients with altered immunity, we over-sampled patients with B-cell/plasma cell malignancies (B-cell) and any cancer patient receiving immune checkpoint inhibitors (ICI). The study was approval by the institutional review board and all participants provided written informed consent.

### Clinical Data Collection

Electronic medical records were reviewed to extract clinical data including tumor type (ICD-10 codes) and anti-cancer treatment regimens and timing of administration relative to vaccination. Active anticancer treatment status was defined as any therapy within six months prior to and up to 14 days post-vaccination. For transplant recipients, patients within one year of autologous stem cell transplant or within two years of allogeneic stem cell transplant from vaccination were regarded as under active treatment. SARS-CoV-2 vaccination status, vaccine type and dates of administration were obtained from medical records and linkage to the California Immunization Registry. We use the date of Dec-25-2021 to define the timepoint when the Omicron SARS-CoV-2 lineage became dominant in the local catchment area. Primer vaccination was defined as receiving either one, two or three doses of mRNA vaccine [Pfizer-BioNtech BNT162b2 (BNT) or Moderna/NIH mRNA-1273 (m1273)] within 120 days, or one-dose of the Janssen Ad26.COV2.S (Ad26). Booster vaccinations were defined as receipt of any type of mRNA vaccine or Ad26 after 120 days the first dose. In August and September 2022, bivalent versions (original and omicron BA.4/BA.5 variant) of the vaccine from Pfizer and Moderna were authorized for use as booster doses in individuals aged 18 years of age or older in the U.S.

### Questionnaire Data

Self-administered questionnaires were sent electronically to patients at the time of enrollment, following vaccinations, 6-month after enrollment and annually thereafter. Surveys included questions regarding medical history, lifestyle, vaccine attitudes and quality of life.

#### Vaccine hesitancy

Perspectives and attitudes towards the vaccine were measured using a modified version of the World Health Organization (WHO) Vaccine Hesitancy Scale and Group-Based Medical Mistrust Scale. The resulting instrument had eight questions and participants were asked to indicate how strongly they agreed or disagreed with a statement using a five-point likert scale.

#### Patient Reported Outcomes (PROs)

The Patient-Reported Outcomes Measurement Information System (PROMIS) short form measures were used to assess anxiety, depression, instrumental support, informational support, and emotional support. Items assessing emotional support inquired about whether respondents had a confidante or someone to talk about problems, or someone who made them feel appreciated.[36] Instrumental support items asked whether respondents had someone to help them take care of daily living tasks as needed, such as helping with chores, running errands, helping out when sick, or helping with transportation for medical care.[36] Items on informational support asked about whether respondents had someone who could give advice, provide suggestions, or give helpful information about life decisions.[36] Higher T scores indicated higher perceived depression, anxiety, emotional, instrumental, and informational social support.

#### Loneliness

The UCLA 3-Item Loneliness Scale [37] was used to capture how often individuals felt that they “lack companionship”, were “left out”, and “isolated from others.” Responses to all items were averaged to create a composite loneliness score with higher values indicating greater loneliness.[38] This scale has been used in previous longitudinal studies and shows good internal consistency, as well as concurrent and discriminant validity.[37]

#### Side effects

A self-administered symptoms questionnaire was given to patients after vaccine administrations (dose 1, 2 and 3) to collect occurrence and severity of injection site pain, headache, fatigue, fever, pain, redness, and/or swelling at the injection site.

### SARS-CoV-2 Immunoglobulin G Antibody Testing

Peripheral blood was collected from participants approximately every 3-6 months. Serological testing for antibodies to the receptor binding domain (RBD) of the S1 subunit of the viral spike protein [IgG(S-RBD)] and antibodies targeting the viral nucleocapsid protein (IgG(N)) was performed using the SARS-CoV-2 IgG II and SARS-CoV-2 IgG assays, respectively [Abbott Diagnostics (Illinois, U.S.)]. We present findings using the WHO unit (binding antibody unit per ml [BAU/ml]) based on the mathematical relationship of BAU/ml=0.142*AU/ml.[39] The minimal threshold for seroconversion was defined as IgG(S-RBD)=7.1 BAU/ml.[40] Detectable antibody responses were classified as “low” (7.1–590.72 BAU/ml) and “high” (≥590.72 BAU/ml). The 590.72 BAU/ml threshold was shown to correspond to a 0.95 probability of obtaining a PRNT ID50 at a 1:250 dilution and is a representative of a high titer and correlate of neutralization.[41] Values were cutoff at 25,000 AU/ml (3,550 BAU/ml).

### T-Cell Receptoire (TCR)

Sequencing of human TCRβ chains was performed on DNA extracted from buffy coat specimens using the immunoSEQ Assay (Adaptive Biotechnologies, Seattle, WA). After bias-controlled multiplex PCR and high-throughput sequencing, the absolute abundance of unique TCRβ CDR3 regions and the corresponding T cell fractions by template count normalization are quantified.[42] SARS-CoV-2 spike or other non-spike SARS-CoV-2 protein specificities for CDR3s are assigned using reference lists generated by Adaptive Biotechnologies, which have been described previously.[29]

The primary metrics used to summarize SARS-CoV-2 TCR results are: T-detect, breadth and depth. T-detect is a binary variable analogous to seropositivity, with a threshold defined by Adaptive Biotechnologies. Clonal breadth is defined as the number of unique annotated rearrangements divided by the total number of unique productive rearrangements in each sample (*1000 in all figures). Clonal depth is calculated by a formula described previously, [43] which can be interpreted as the relative number of clonal expansion generations across the TCRs, as normalized by the total number of TCRs in the sample.[44]

### Statistical Analysis

Time of vaccine uptake was visualized using cumulative incidence curve and compared by age and self-reported race/ethnicity categories using log-rank tests. Patient demographic and clinical characteristics were summarized by cancer type (solid v. hematologic) using median and interquartile range (IQR) or mean and standard deviation (SD) for continuous variables and frequencies and percentages for categorical variables. Kruskal-Wallis or Wilcoxon Rank Sum tests were used to compare continuous variables and Chi-squared tests or Fisher’s exact tests for categorical variables. Peak quantitative IgG(S-RBD) levels were compared across patients’ characteristics using Kruskal-Wallis test at time of priming and boosting separately. Similarly, TCR depth and breadth were compared across patient characteristics at vaccine timepoints. The correlation between IgG levels and TCR depth and breadth were examined by calculating Spearman’s rank correlation coefficient.

For PROs, scores were converted to a T-score, which is used to compare sample scores versus the general population (mean=50, SD=10). Higher T-scores correspond to higher levels (for example, greater anxiety). For the loneliness scale, scores were summed and ranged from 3 (least) to 9 (most lonely). Mixed models and least-squares means were used to compare constructs over the time course of vaccine recommendations adjusted for infection and status of vaccine uptake.

Primary clinical outcomes of interest were SARS-CoV-2 infection (at any timepoint) and all-cause mortality. Cox models with time dependent covariates were used to examine associations of selected predictors and SARS-CoV-2 infection and all-cause mortality. We focused on the following demographic and clinical predictors: age, sex, race and ethnicity, tumor type, treatment, ECOG status, booster vaccination status and infection statu. For all-cause mortality, we excluded individuals who were never vaccinated (76 individuals) and who died within 120 days from the first vaccination (9 individuals).

*P* value <.05 was considered statistically significant. Statistical analysis was performed in R, version 4.2.2.

## Data Availability

The current study is funded by NCI SeroNet U54 and as a requirement data is made publicly available via ImmPort.

## Acknowledgements

We sincerely appreciate all SeroNet-CORALE participants and study staff. This work was supported by NCI SeroNet U54 CA260560, and by the Biobank and Biostatistics Shared Resource of Cedars-Sinai Cancer. We are also grateful to all the front-line healthcare workers in our healthcare system who continue to be dedicated to delivering the highest quality care for all patients.

## Authors Contributions

J.C.F, A.M., K.R., N.M. were responsible for the conceptualization. K.R., R.V., J.D., J.G., N.N., A.G. W.T., M.N., J.L., N.M., O.M., R.N., I.M., A.H., A.M., R.P., R.N. were responsible for patient recruitment. N.N., K.S., W.T., S.C. were responsible for the sample preparation. S.S. and J.C.F. were responsible for the selection of surveys. S.C., J.C.F., E.K., K.S., T.L., S.M., O.H. were responsible for the data analysis and data curation. J.C.F, A.M., S.C., N.M., O.H. were responsible for interpreting findings. J.C.F., K.R., A.M. W.T., N.M., O.H. were responsible for the overall supervision and funding acquisition. This work was supported by the Biobank and Biostatistics Shared Resources of Cedars-Sinai Cancer (S.C., V.K.R., C.H., W.T., and K.S.) All authors were responsible for reviewing and editing the final manuscript.

## Inclusion and Diversity

We support inclusive, diverse and equitable conduct of research.

## Supplementary Appendix

**Figure S1:**
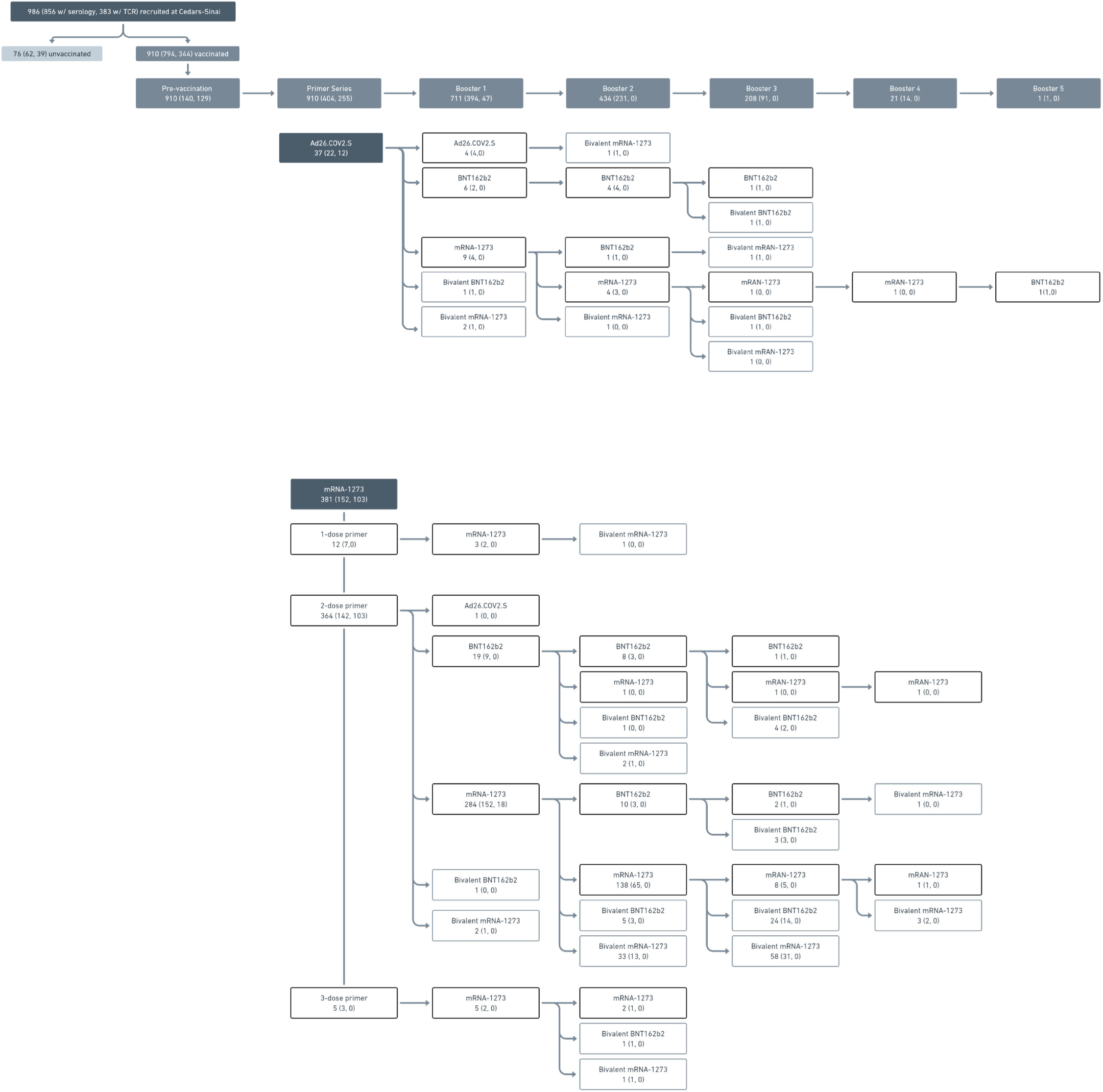
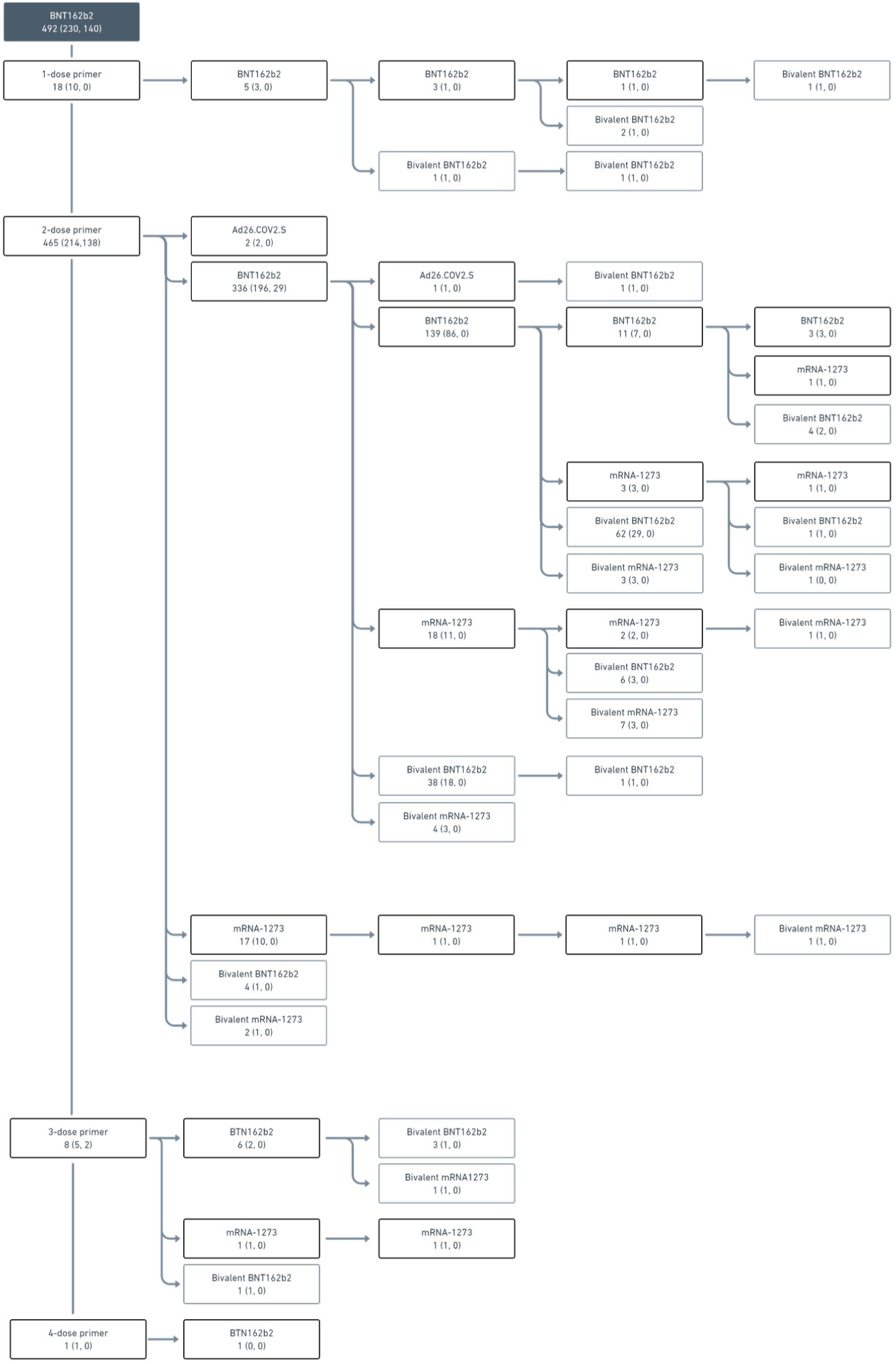
Consort diagram of study participants and their vaccine status

**Figure S2:**
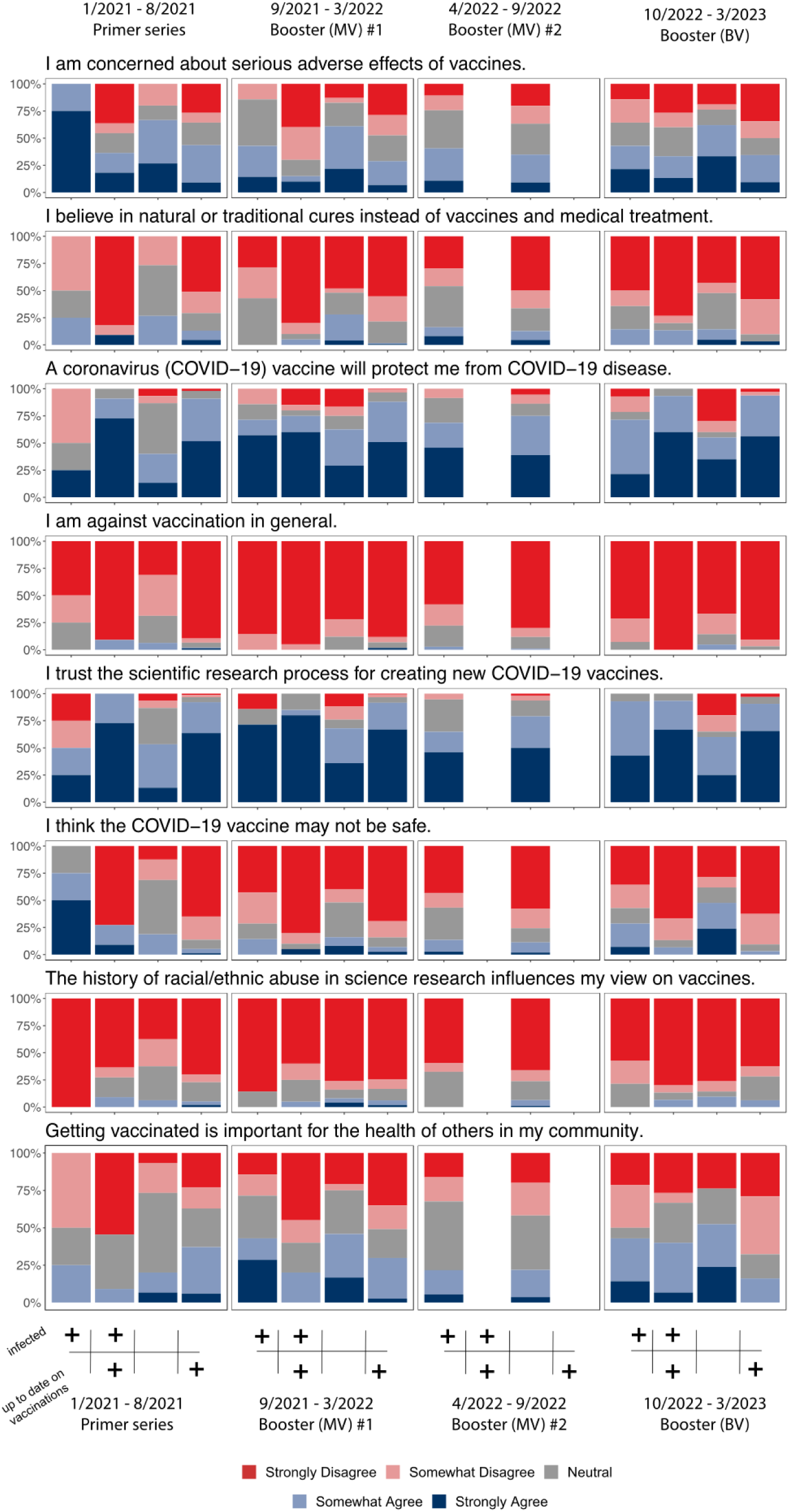
Vaccine perspectives among patients with cancer

**Figure S3:**
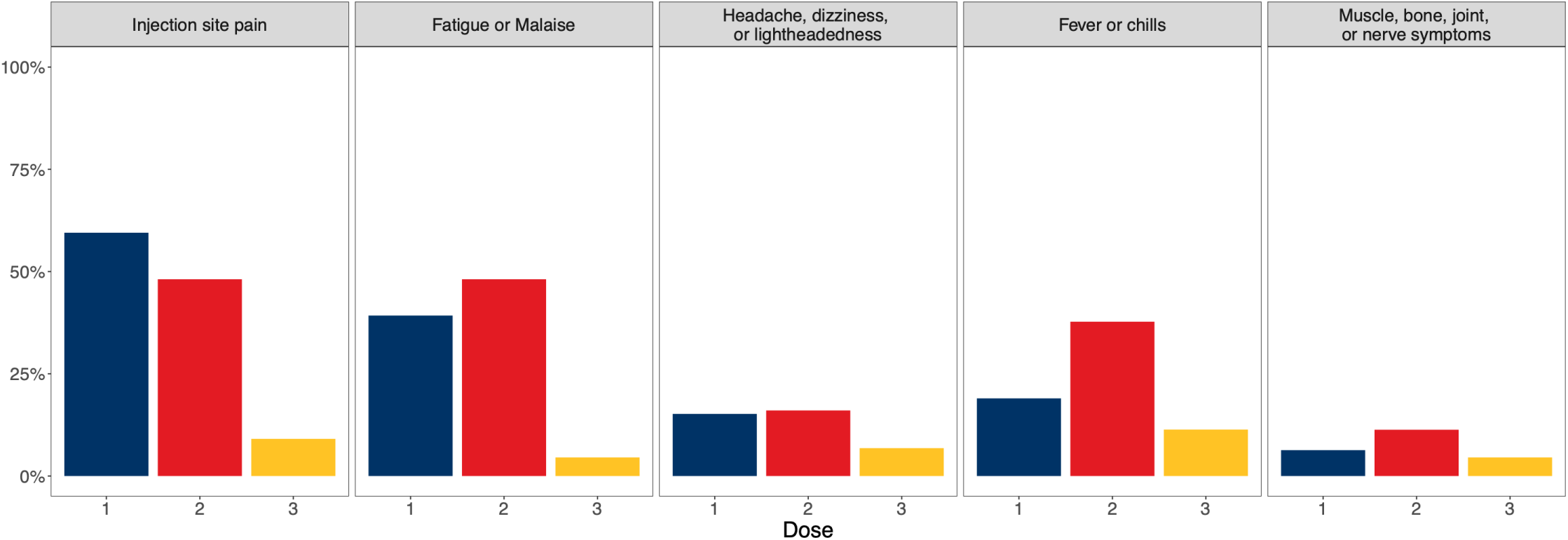
Acute side-effects after vaccination among cancer patients

**Figure S4:**
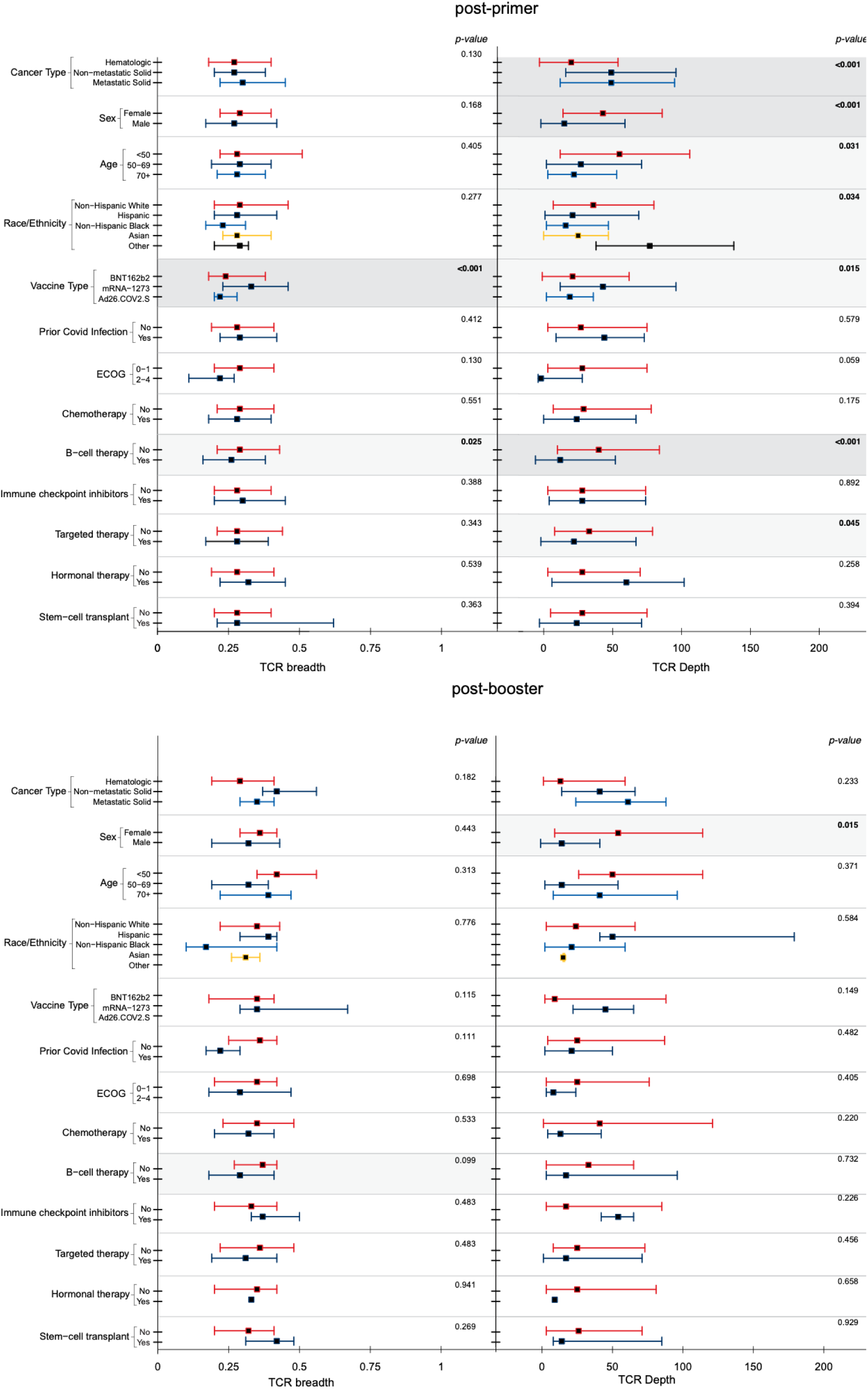
Post-vaccination T-cell repertoire responses among cancer patients

**Table S1:**
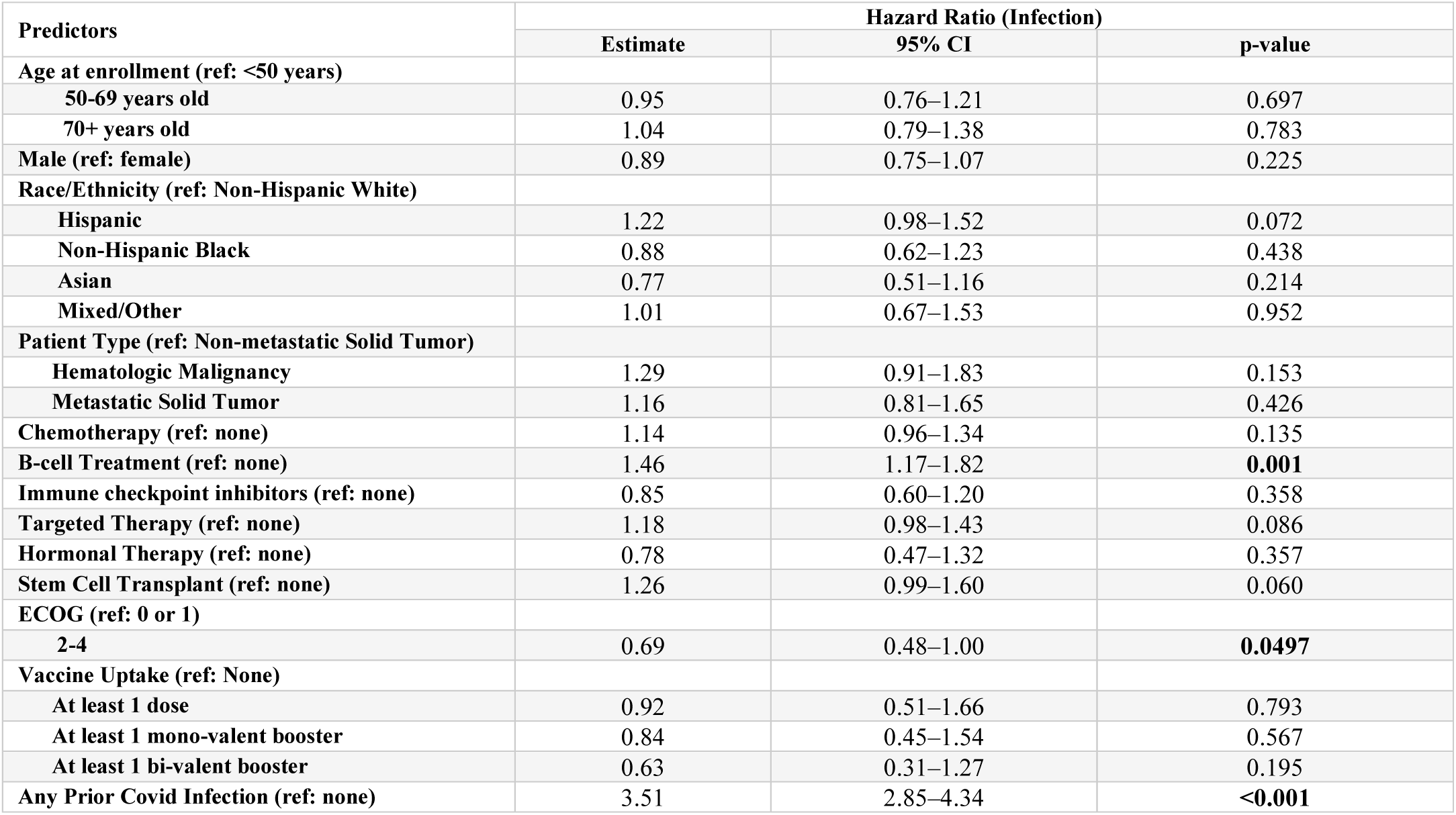
Multivariable analyses of risk of SARS-CoV-2 infection among patients with cancer.

| Predictors | Hazard Ratio (Infection) |  |  |
| --- | --- | --- | --- |
|  | Estimate | 95% CI | p-value |
| Age at enrollment (ref: <50 years) |  |  |  |
| 50-69 years old | 0.95 | 0.76–1.21 | 0.697 |
| 70+ years old | 1.04 | 0.79–1.38 | 0.783 |
| Male (ref: female) | 0.89 | 0.75–1.07 | 0.225 |
| Race/Ethnicity (ref: Non-Hispanic White) |  |  |  |
| Hispanic | 1.22 | 0.98–1.52 | 0.072 |
| Non-Hispanic Black | 0.88 | 0.62–1.23 | 0.438 |
| Asian | 0.77 | 0.51–1.16 | 0.214 |
| Mixed/Other | 1.01 | 0.67–1.53 | 0.952 |
| Patient Type (ref: Non-metastatic Solid Tumor) |  |  |  |
| Hematologic Malignancy | 1.29 | 0.91–1.83 | 0.153 |
| Metastatic Solid Tumor | 1.16 | 0.81–1.65 | 0.426 |
| Chemotherapy (ref: none) | 1.14 | 0.96–1.34 | 0.135 |
| B-cell Treatment (ref: none) | 1.46 | 1.17–1.82 | **0.001** |
| Immune checkpoint inhibitors (ref: none) | 0.85 | 0.60–1.20 | 0.358 |
| Targeted Therapy (ref: none) | 1.18 | 0.98–1.43 | 0.086 |
| Hormonal Therapy (ref: none) | 0.78 | 0.47–1.32 | 0.357 |
| Stem Cell Transplant (ref: none) | 1.26 | 0.99–1.60 | 0.060 |
| ECOG (ref: 0 or 1) |  |  |  |
| 2-4 | 0.69 | 0.48–1.00 | **0.0497** |
| Vaccine Uptake (ref: None) |  |  |  |
| At least 1 dose | 0.92 | 0.51–1.66 | 0.793 |
| At least 1 mono-valent booster | 0.84 | 0.45–1.54 | 0.567 |
| At least 1 bi-valent booster | 0.63 | 0.31–1.27 | 0.195 |
| Any Prior Covid Infection (ref: none) | 3.51 | 2.85–4.34 | **<0.001** |

